# Differentiation of SARS-CoV-2 naturally infected and vaccinated individuals in an inner-city emergency department

**DOI:** 10.1101/2021.10.13.21264968

**Authors:** Evan J Beck, Yu-Hsiang Hsieh, Reinaldo E Fernandez, Gaby Dashler, Emily R Egbert, Shawn A Truelove, Caroline Garliss, Richard Wang, Evan M. Bloch, Ruchee Shrestha, Joel Blankson, Andrea L. Cox, Yukari C Manabe, Thomas Kickler, Richard E Rothman, Andrew D Redd, Aaron AR Tobian, Aaron M Milstone, Thomas C Quinn, Oliver Laeyendecker

## Abstract

**Background:** Emergency Departments (EDs) can serve as surveillance sites for infectious diseases. Our purpose was to determine the burden of SARS-CoV-2 infection and prevalence of vaccination against COVID-19 among patients attending an urban ED in Baltimore City.

**Methods:** Using 1914 samples of known exposure status, we developed an algorithm to differentiate previously infected, vaccinated, and unexposed individuals using a combination of antibody assays. We applied this testing algorithm to 4360 samples ED patients obtained in the springs of 2020 and 2021. Using multinomial logistic regression, we determined factors associated with infection and vaccination.

**Results:** For the algorithm, sensitivity and specificity for identifying vaccinated individuals was 100% and 99%, respectively, and 84% and 100% for naturally infected individuals. Among the ED subjects, seroprevalence to SARS-CoV-2 increased from 2% to 24% between April 2020 and March 2021. Vaccination prevalence rose to 11% by mid-March 2021. Marked differences in burden of disease and vaccination coverage were seen by sex, race, and ethnicity. Hispanic patients, though 7% of the study population, had the highest relative burden of disease (17% of total infections) but similar vaccination rates. Women and White individuals were more likely to be vaccinated than men or Black individuals (adjusted odds ratios [aOR] 1.35 [95% CI: 1.02, 1.80] and aOR 2.26 [95% CI: 1.67, 3.07], respectively).

**Conclusions:** Individuals previously infected with SARS-CoV-2 can be differentiated from vaccinated individuals using a serologic testing algorithm. SARS-CoV-2 exposure and vaccination uptake frequencies reflect gender, race and ethnic health disparities in this urban context.

**Summary:** Using an antibody testing algorithm, we distinguished between immune responses from SARS-CoV-2-infected and vaccinated individuals. When applied to blood samples from an emergency department in Baltimore, disparities in disease burden and vaccine uptake by sex, race, and ethnicity were identified.

## Introduction

As of October 2021, over 238 million cases of Severe Acute Respiratory Syndrome Coronavirus 2 (SARS-CoV-2) infection, which causes coronavirus disease 2019 (COVID-19), have been reported globally^1^. The United States has recorded more than 700,000 deaths and documented infections in over 10% of the population. Within the U.S., Black and Latino individuals have experienced higher rates of infection and mortality, relative to White individuals, since the onset of the pandemic^2,3^. Rooted in long-standing racial and structural injustice, these disparate health outcomes reflect the disproportionate effects of social determinants of health among U.S. racial and ethnic minority groups^4,5^.

Currently, three vaccines for COVID-19 have been authorization by the U.S. Food and Drug Administration^6^. The authorized vaccines from Pfizer, Moderna, and Johnson & Johnson, each elicit an immune response against the spike protein of the SARS-CoV-2 virion^7–9^. As of the 20^th^ of September 2021, 74.6% of persons aged ≥ 12 years have received at least one dose of a COVID-19 vaccine in the United States^10^. This uptake, however, has varied dramatically by race, socioeconomic status, and geographic location. Several studies have described the potential for vaccine hesitancy among Black and Hispanic Americans^11–13^. These issues related vaccine hesitancy and access could result in disparate rates of vaccine uptake among racial and ethnic minority groups.

In contrast to vaccinated individuals, naturally infected patients create antibodies to several parts of the virus, including the spike and nucleocapsid proteins^14^. By comparing the results of serologic assays that either detect antibodies to spike (S1), the spike glycoprotein receptor binding domain (RBD), or the nucleocapsid (N), it should be possible to distinguish between SARS-CoV-2 naturally infected (either infected alone or infected then vaccinated), vaccinated (with no evidence of prior infection), and uninfected individuals.

By providing data on symptomatic infection rates, emergency departments (EDs) have historically played a critical role in prior epidemics and pandemics and thus present a rich opportunity for conducting SARS-CoV-2 serosurveillance^15–17^. Although case-reporting can provide an estimate of population-level seroprevalence, relying on case-reporting alone may underestimate the burden of infection, emphasizing the need for accurate serologic assessment of seroprevalence^18^.

## Methods

### Ethics Statement

This study used samples from parent studies approved by The Johns Hopkins University School of Medicine Institutional Review Board (IRB00245545, IRB00247886, IRB00091667, IRB00250798, IRB00249350, and NA_00085477). The Moderna vaccine trial was part of the Division of Microbiology and Infectious Diseases Protocol Number: 20-0003. For those studies, all individuals provided written informed consent. The JHU School of Medicine Institutional Review Board (IRB00083646, CIR00016268) approved the de-identified serosurvey performed on waste material. All studies were conducted according to the ethics standards of the Helsinki Declaration of the World Medical Association.

### Samples for Algorithm Validation

Three sample sets with known previous infection and vaccination to SAR-CoV-2 were used to validate the antibody testing algorithm (**Table 1**). Samples with known vaccination were drawn from a phase I trial^8^ (n=68) and vaccinated health care professionals (HCP, n=360)^20,21^. The 494 samples from individuals known to have been infected by SARS-CoV-2 were drawn from three cohorts: convalescent plasma donors (CCP, n=244)^18,22^; and Clinical Characterization Protocol for Severe Infectious Diseases (CCPSEI, n=246)^23^, and HCP (n=4)^24^. All samples were from individuals with a known positive SARS-CoV-2 RT-PCR test result. The majority of the CCP donors had mild disease, with 9% of this cohort reporting hospitalization. Among the CCPSEI, 14% received oxygen therapy, 33% received ventilation, and 13% died. All HCP had mild disease. Additionally, there were 46 HCP who were infected and subsequently vaccinated, 28 with confirmed a PCR date and 18 who were suspected as having prior infection (PCR negative, but symptomology indicative of an infection). Specificity of the testing algorithm was then assessed using 992 samples from pre-pandemic remnant CBC samples collected from Johns Hopkins Hospital Emergency Department (JHH ED) patients collected between December 2015 and January 2016^25^.

**Table 1:** Description of Cohorts

| Cohort Name<br>(IRB number) | Purpose | Number of<br>Samples | Notes |
| --- | --- | --- | --- |
| NIH Phase 1 Vaccine<br>Trial (20-0003) | Positive Control –<br>Known SARS-CoV-2 Vaccinated | 68 | 68 vaccinated samples with no<br>prior infection |
| JHHS Health Care<br>Professionals<br>(IRB00249350) | Positive Control –<br>Known SARS-CoV-2 Vaccinated<br>& some naturally<br>infected | 410 | 360 vaccinated samples with no<br>prior infection<br>28 vaccinated samples with<br>prior infection<br>18 vaccinated samples with<br>suspected prior infection<br>4 unvaccinated sample with<br>prior infection |
| Potential Convalescent<br>Plasma Donors<br>(IRB00250798) | Positive Control –<br>Known SARS-CoV-2 Infected | 244 | Known infected (PCR positive)<br>and non-vaccinated |
| CCPSEI<br>(IRB00247886 and<br>IRB00091667) | Positive Control –<br>Known SARS-CoV-2 Infected | 246 | Known infected (PCR positive)<br>and non-vaccinated |
| JHH ED 2016<br>(NA_00085477) | Pre-Pandemic<br>negative control | 992 | Pre-pandemic samples from the<br>same survey site as pandemic<br>surveillance site |
| JHH ED 2020: 16 March<br>to 30 April<br>(IRB00083646,<br>CIR00016268) | Algorithm<br>Application | 1536 | Population surveillance |
| JHH ED 2021: 11<br>January to 10 March<br>(IRB00083646,<br>CIR00016268) | Algorithm<br>Application | 2824 | Population surveillance |

Abbreviations: CCPSEI, Clinical Characterization Protocol for Severe Infectious Diseases; ED, emergency department; JHHS, Johns Hopkins Healthcare System; JHH Johns Hopkins Hospital; PCR, polymerase chain reaction; SARS-CoV-2, Severe Acute Respiratory Syndrome Coronavirus 2.

### Samples for Algorithm Application

The testing algorithm was subsequently applied to two serosurveys conducted among patients attending the JHH ED from 16 March to 30 April 2020 and from 11 January to 10 March 2021. As previously described in an identity-unlinked seroprevalence study^26^, remnant CBC blood samples from ED patients aged >17 years were collected during the study period. Each sample was assigned a unique study code, processed, and stored at −80°C. Basic patient demographic characteristics (age, sex, race, and ethnicity) were abstracted from the ED administrative database, and all identifiers and protected health information removed from the dataset. Data regarding COVID-19 vaccination status was not available. Laboratory testing was then performed on stored specimens after delinking the demographic dataset. Using the unique study code, SARS-CoV-2 serostatus and demographic data were then merged.

### Laboratory Methods

The testing algorithm required three serologic assays that could differentiate serologic reactivity to SARS-CoV-2 S1, RBD and nucleocapsid. These assays were limited to standard ELISA and point of care assays, as we did not have access to chemiluminescent detection equipment. We selected ELISA based technologies for the initial high throughput screening, thought confirmation testing could include point of care assays. Additional information on the assays used is available in **Supplemental Table 1**.

Plasma and serum samples were analyzed using three commercially available serologic assays: the Euroimmun Anti-SARS-CoV-2 ELISA (Mountain Lakes, NJ), the CoronaCHEK^™^ COVID-19 IgG/IgM Rapid Test Cassette (Hangzhou Biotest Biotech Co Ltd), and the Bio-Rad Platelia SARS-CoV-2 Total Antibody ELISA (Marnes-la-Coquette, France). Each assay was selected for previously determined performance^27,28^, ease-of-use characteristics (standard ELISA technology, no large pieces of equipment necessary) and availability. The Euroimmun ELISA measures IgG responses to the S1 protein of SARS-CoV-2, whereas the Bio-Rad ELISA measures total antibodies to nucleocapsid. Both ELISA assays generate a ratio of the optical density of the sample divided by the control (referred to as a signal to cut-off ratio [S/C]). For the Euroimmun and Bio-Rad ELISAs, an S/C ≥ 0.8 was considered a positive result. The CoronaCHEK^™^ lateral flow assay (LFA) tests for the presence of both IgM and IgG antibodies to the receptor-binding domain (RBD) of the spike protein. Any visual band was considered a positive result. Each assay was performed according to the manufacturer’s instructions.

An algorithm composed of the Euroimmun, Bio-Rad, and CoronaCHEK assays was used to differentiate samples into three groups: naturally infected (who may or may not subsequently be vaccinated); vaccinated (who were never infected); and unexposed (**Figure 1)**. All samples were first tested using the Euroimmun ELISA (S1). Next, all positive and indeterminate samples were subsequently tested on CoronaCHEK (RBD). Samples that tested positive on Euroimmun and negative on CoronaCHEK were assumed to be false positives and classified as not naturally infected or vaccinated (unexposed). Samples that tested positive on CoronaCHEK were then tested with the Bio-Rad Total Ab assay (N). Samples which were reactive by Euroimmun and CoronaCHEK but negative for Bio-Rad were considered vaccinated. Those samples with a positive or indeterminate result on Bio-Rad were considered natural infections.

**Figure 1.**
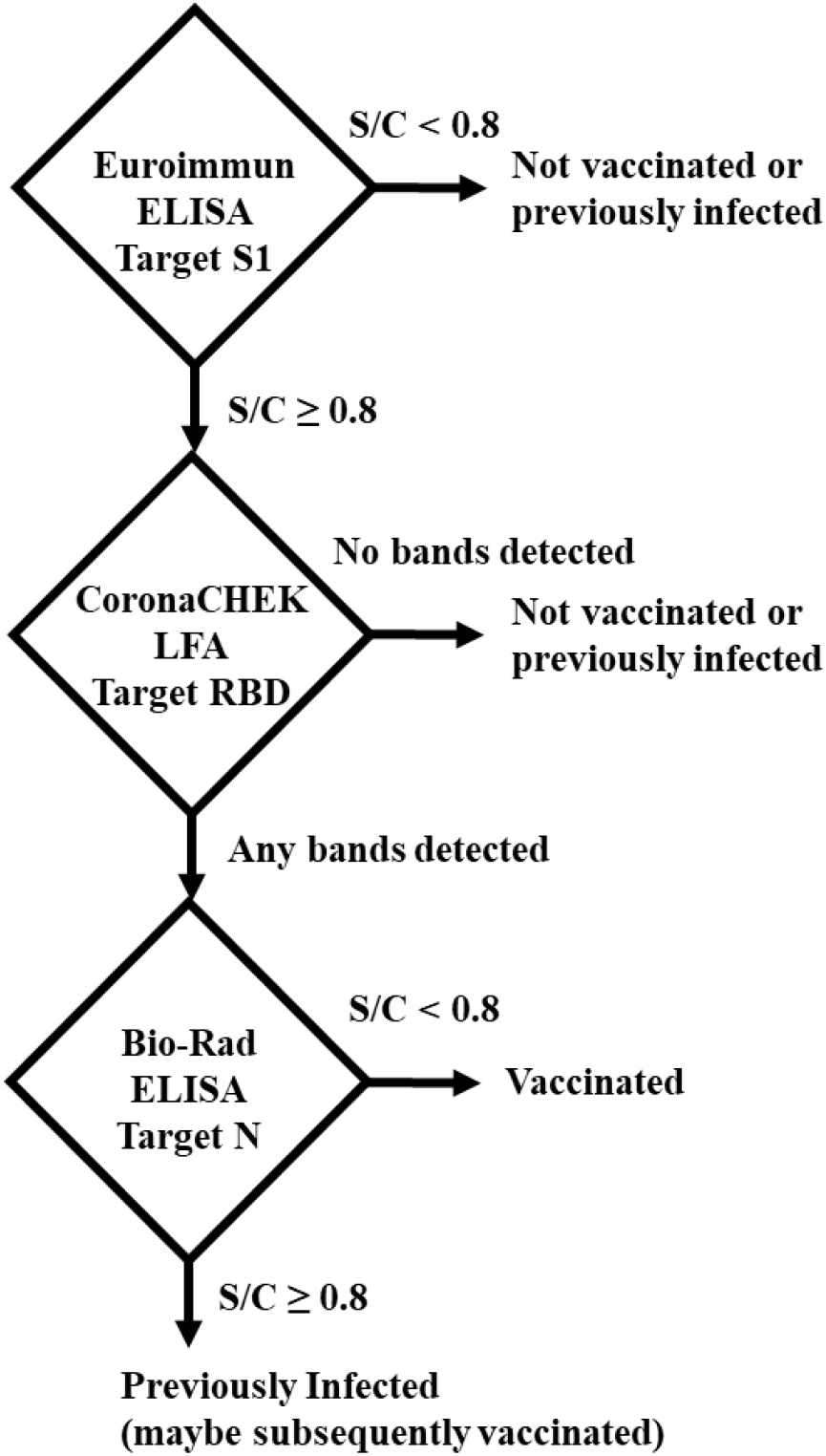
Antibody testing algorithm. An S/C ≥ 0.8 result on the Euroimmun Anti-SARS-CoV-2 ELISA (S-1) and positive result on the CoronaCHEK^™^ COVID-19 IgG/IgM Rapid Test Cassette (RBD) were considered positive for the SARS-CoV-2 spike protein. An S/C ≥ 0.8 result on Bio-Rad Platelia SARS-CoV-2 Total Antibody ELISA (nucleocapsid) was considered a natural infection, whereas an S/C < 0.8 in combination with a positive result for spike/RBD indicated vaccination. Samples with negative tests on either Euroimmun or CoronaChek were considered unexposed to either SARS-CoV-2 infection or COVD-19 vaccination.

### Statistical methods

To evaluate the diagnostic accuracy of the testing algorithm for a particular state (vaccinated or naturally infected), the sensitivity for each state was determined from samples with that known status (the training sample sets; **Table 1**). Calculation of specificity included all samples from unexposed individuals and the samples from individuals of the other state. Since sample collection had occurred prior to the availability of the COVID-19 vaccines, both naturally infected and pre-pandemic samples were considered negative samples to calculate the sensitivity and specificity of the algorithm for the detection of vaccinated samples. To calculate algorithm sensitivity and specificity for naturally infected samples, samples from the vaccinated (known to be uninfected) and pre-pandemic cohorts were considered negative samples. The 48 samples from individuals known or suspected to be infected and subsequently vaccinated were not include in determining the performance of the algorithm. Statistically significant differences in the ELISA S/C values between vaccinated and naturally infected individuals were determined using a t-test. Chi-squared and Fisher’s exact tests was used to examine the differences in population demographics between the 2020 and 2021 serosurveys. For the JHH ED serosurvey sample sets, factors associated with natural infection or vaccination were assessed with logistic regression. Logistic regression was used to generate odds ratios to compare the odds of natural infection and vaccination in the post-vaccine era with respect to age, sex, and race in the post-vaccine era (spring 2021). A p-value <0.05 was considered significant. Statistical analyses were performed using StataSE version 14.2 (StataCorp, College Station, TX) and SAS version 9.4 (SAS Institute., Cary, NC).

## Results

Samples from vaccinated individuals without prior SARS-CoV-2 infection (n=428), unvaccinated individuals with PCR confirmed natural infection (n=494), and those seen in the ED pre-pandemic (n=992) were tested on all three assays (**Supplemental Figure 1a**). The Euroimmun S1 ELISA was positive for 100% (95% confidence interval [CI], 99.1-100.0%), 89% (95% CI, 86.2-91.9%), and 3.2% (95% CI, 2.2-4.5%) for vaccinated, naturally infected and pre-pandemic samples, respectively. Similarly, for the Bio-Rad N ELISA, 0% (95% CI, 0.0-0.7%), 91% (95% CI, 88.2-93.5%), and 1.4% (95% CI, 0.7-2.4%) were positive for vaccinated, naturally infected and pre-pandemic sample sets. For the CoronaCHEK RBD assay, 100% (95% CI, 99.1-100.0%), 91% (95% CI, 87.8-93.1%), and 0.5% (95% CI, 0.2-1.2%) had any reactive band for vaccinated, naturally infected and pre-pandemic samples. For vaccinated samples, algorithm sensitivity and specificity were 100% (95% CI, 99.1-100.0%) and 98.9% (95 Cl, 98.2-99.3%), respectively (**Supplemental Figure 1b**). For naturally infected samples, algorithm sensitivity and specificity were 84.4% (95% CI, 80.9-87.5%) and 100% (95% CI, 99.7-100.0%), respectively (**Supplemental Figure 1c**).

There were significant differences between SARS-CoV-2 serostatus and the level of antibody reactivity to spike and nucleocapsid among the cohorts used for algorithm validation (**Figure 2**). For vaccinated individuals, the median S/C value for antibody reactivity against spike was 8.9 (IQR=1.2) compared to 5.2 (IQR=5.3) for infected persons (p<0.001). Among the vaccinated persons without previous infection, no individuals had an S/C value for antibody reactivity against nucleocapsid greater than 0.8, the threshold for a positive result, whereas naturally infected patients with no history of vaccination had a median S/C of 4.3 (IQR=0.53) (p<0.001). Among HCP, there were 28 samples from individuals with a known SARS-CoV-2 PCR positive date who were vaccinated 7 to 103 days later. These individuals had spike antibody S/C values similar to vaccinated individuals (median=9.5, IQR=0.9) and nucleocapsid antibody S/C values similar to naturally infected individuals (median=3.4, IQR=2.9). Additionally, 18 HCP who were SARS-CoV-2 PCR negative but had suspected infection, had similar values to those with known infection followed by vaccination, with spike antibody levels (median=10.0, IQR=1.2) and nucleocapsid antibody S/C values (median=3.3 IQR 1.9). Because of the timing of sample collection relative to vaccination, it is very unlikely that these 18 samples represented breakthrough infections. It is more likely that these infections were unconfirmed infections that occurred prior to vaccination. There was little reactivity for samples from pre-pandemic samples.

**Figure 2.**
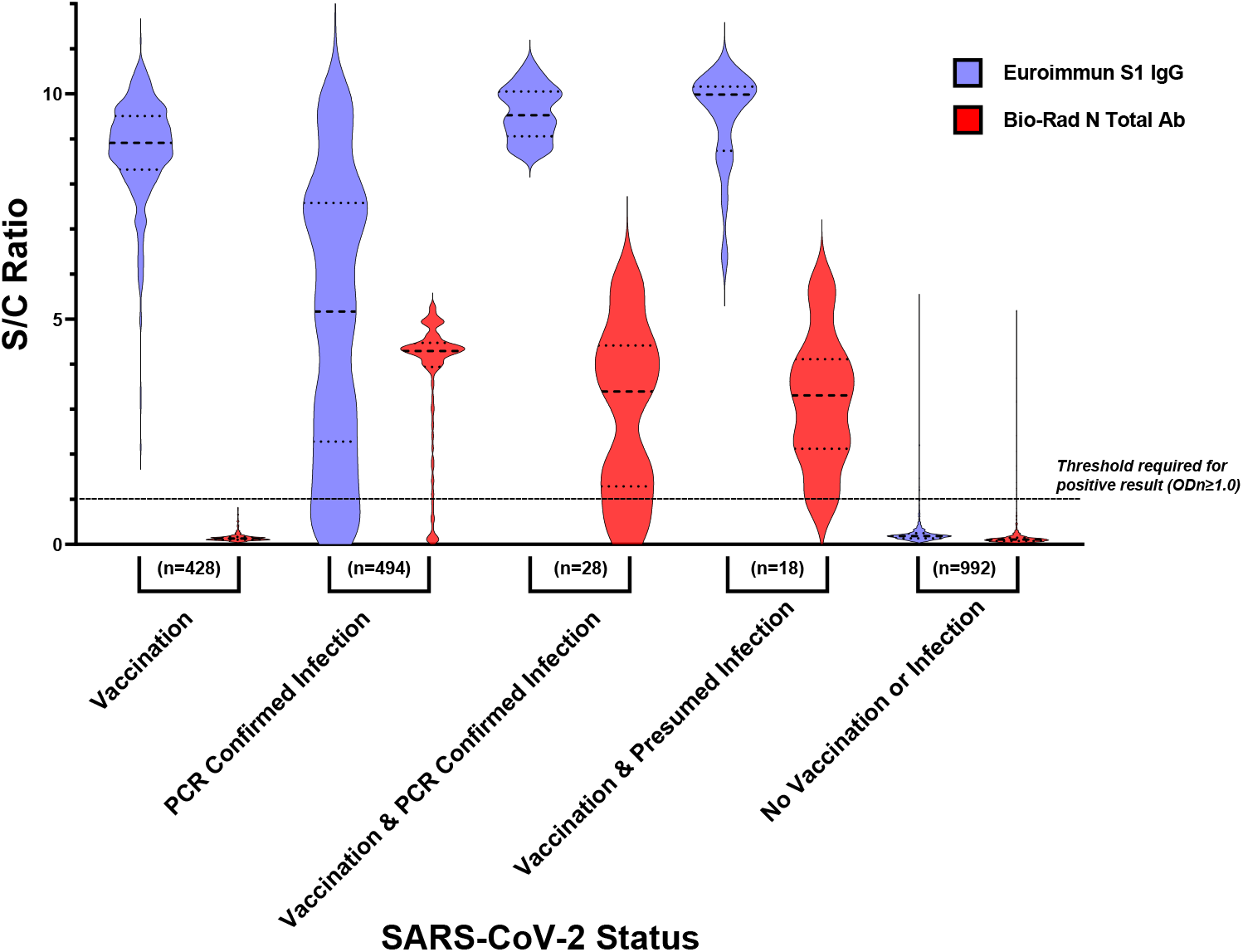
Comparison of ELISA values between vaccinated and naturally infected individuals. Samples with known serostatus from the algorithm validation cohorts were tested on both the Euroimmun Anti-SARS-CoV-2 IgG ELISA (spike) and on Bio-Rad Platelia SARS-CoV-2 Total Antibody ELISA (nucleocapsid). Each ELISA assay generates a ratio of the optical density of the sample divided by a manufacturer-provided calibrator. The y-axis is given as a signal to cut- off ratio (S/C). Medians and interquartile ranges are displayed for each violin plot. The vaccinated group was comprised of individuals with documented vaccination and no previous positive PCR or serological result. SARS-CoV-2 infections were confirmed by a positive PCR result. The vaccination and confirmed infection group was composed of individuals with both documented vaccination and PCR positive infection. Presumed infections were characterized by a lack of PCR positive result, but a positive result for nucleocapsid on the Bio-Rad assay. Samples in the not vaccinated or infected category were obtained from the JHH ED in 2016, prior to the advent of the COVID-19 pandemic.

Using the testing algorithm, 1536 JHH ED 2020 samples and 2824 JHH ED 2021 samples were evaluated. During the two collection periods, combined seroprevalence of antibodies to SARS-CoV-2 from 1.6% (95% CI, 1.1-2.5%) to 23.8% (95% CI, 22.2-25.4%) (**Figure 3**). During the seven weeks of the second survey, the prevalence of vaccination significantly increased from 2.8% (95% CI, 0.9-6.3%) in mid-January to 11% (95% CI, 8.6-13.7%) by mid-March 2021. The age, sex, and race/ethnic demographics of the two survey periods were similar (**Table 2**). For both surveys, approximately 27% were ≥60 years of age, 52% female, 60% Black, 26% White and 7% Hispanic. The prevalence of infection in the spring of 2020 did not vary significantly by age, ethnicity, race and sex. In contrast, by the spring of 2021, significant differences in infection by age ethnicity, race and sex were observed.

**Table 2:**
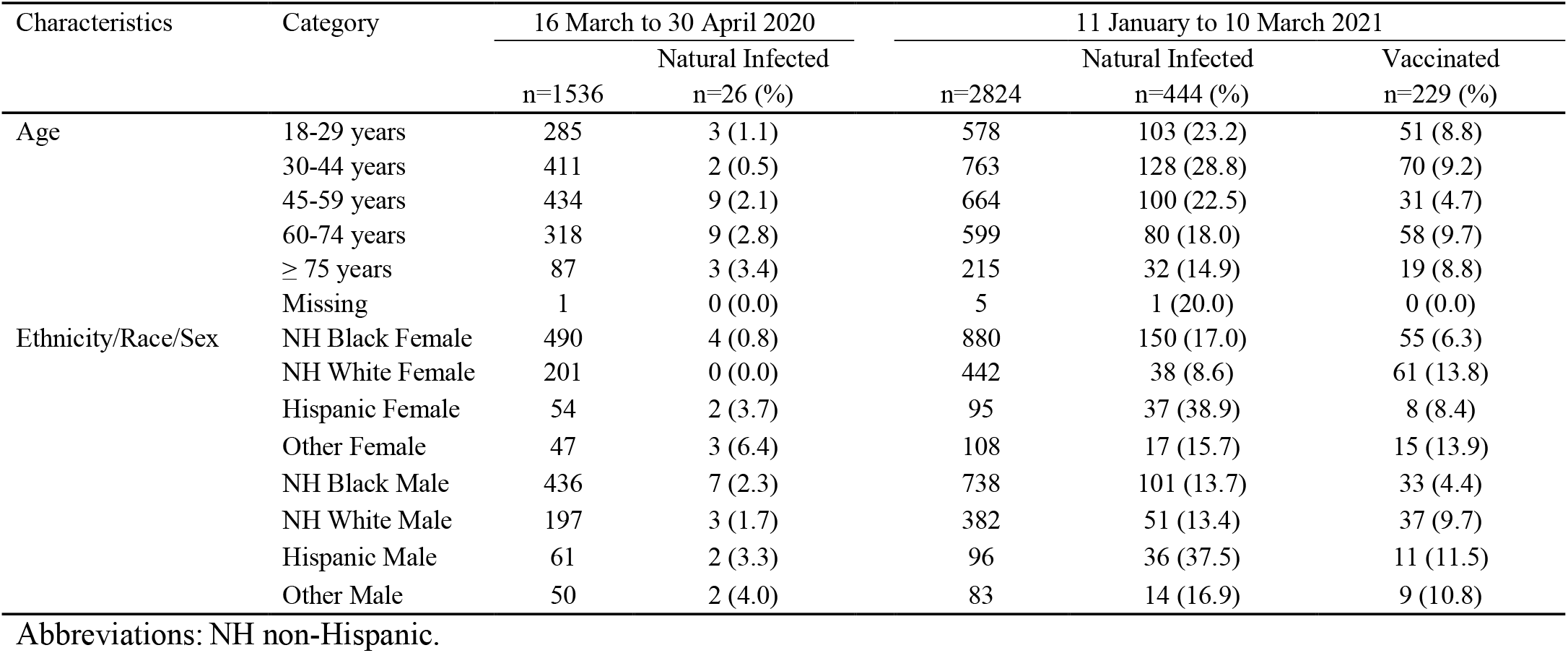
Demographic Characteristics and Seroprevalence of Infection and Vaccination in Emergency Department Patients in the Spring of 2020-2021

**Figure 3.**
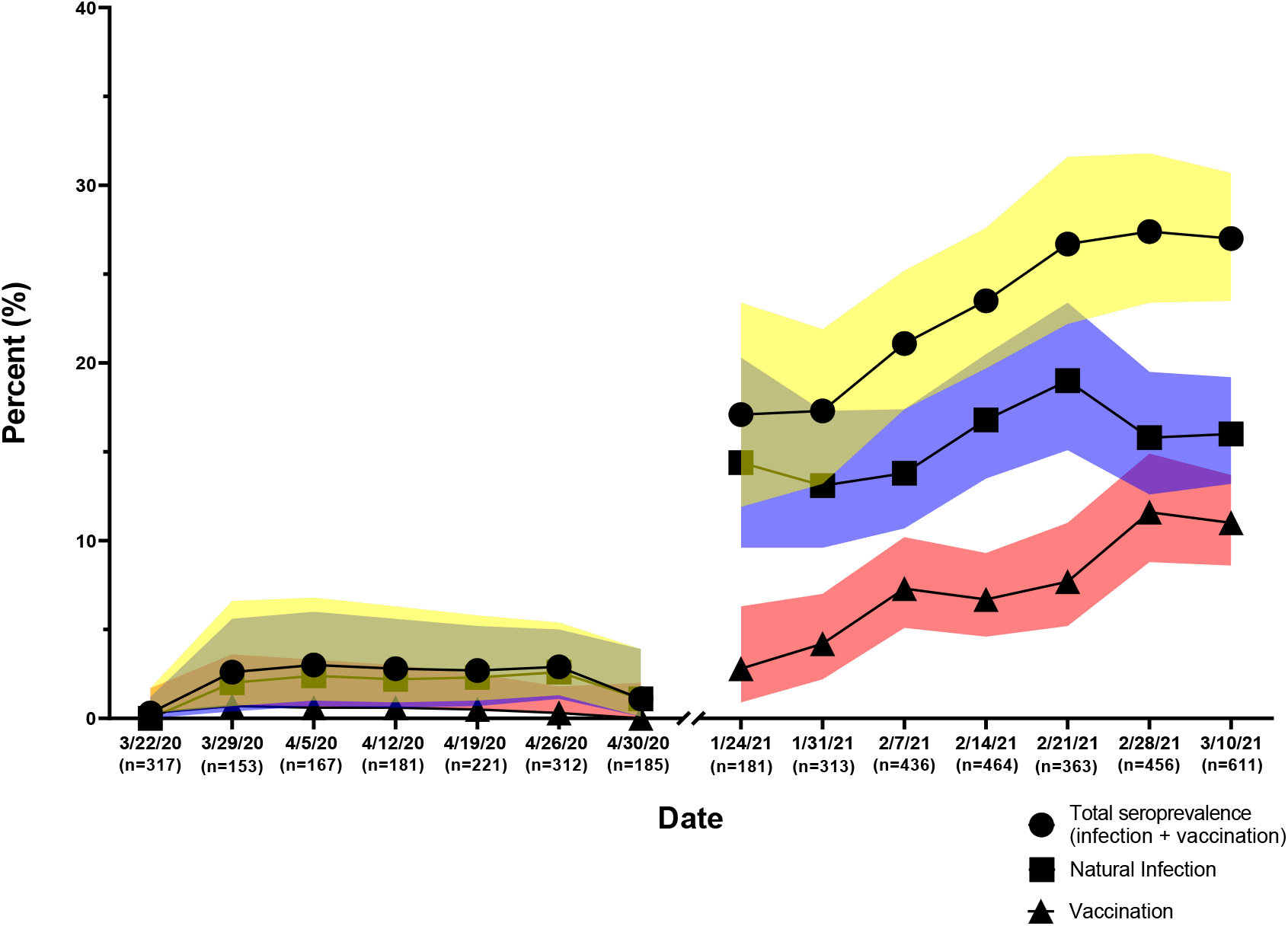
Seroprevalence of antibodies of SARS-CoV-2 2020-2021. JHHED ED samples from 2020 and 2021 were tested on the previously mentioned algorithm and categorized according to the date on which the sample was drawn.

The prevalence of antibodies to SARS-CoV-2 indicating previous infection or vaccination is presented in **Table 2**. White women and men had the lowest prevalence of infection both in 2020 and 2021. White women were the only group that had a higher proportion of vaccinated individuals (14%) compared to infected individuals (9%). In the 2021 survey, White women comprised 9% of all infections in 2021, but 27% of all vaccinations. For all other groups, the prevalence of exposure to SARS-CoV-2 was higher than the frequency of vaccination. By the spring of 2021, Hispanic patients had the highest evidence of prior SARS-CoV-2 infection within any ethnic group at 38%.

In the 2021 survey, there were no statistically different rates of infection between age groups (Table 3). In contrast, 45- to 59-year-olds were less likely to be vaccinated compared to the youngest individuals (aOR=0.71 (95% CI 0.52, 0.98). Compared to Black women, White women were less likely to be previously infected, aOR 0.46 (95% CI 0.31, 0.67), while Hispanic women and men were three times as likely to be previously infected, aOR 3.11 (95% CI 1.98, 4.86) and 2.92 (1.86, 4.58) respectively. When it came vaccination status, White women and men and Hispanic men were all significantly more likely than Black women to have evidence of vaccination, aOR 2.42(95% CI 1.64, 3.56), 1.59 (1.02, 2.47) and 2.04 (95%CI 1.02, 4.08) respectively.

**Table 3.** Factors associated with Antibody Positive to SARS-CoV-2 Among Individuals Attending the JHH ED between 11 January to 10 March 2021.

| Characteristics | Category | OR of Natural Infection<br>(95% CI) | Adj. OR of Natural Infection<br>(95% CI) | OR of Vaccination<br>(95% CI) | Adj. OR of Vaccination<br>(95% CI) |
| --- | --- | --- | --- | --- | --- |
| Age | 18-29 years | 1 | 1 | 1 | 1 |
|  | 30-44 years | 0.93 (0.70, 1.24) | 0.86 (0.64, 1.16) | 1.04 (0.72, 1.52) | 0.99 (0.67, 1.46) |
|  | 45-59 years | 0.82 (0.61, 1.11) | 0.84 (0.62, 1.14) | **0.51 (0.32, 0.80)** | **0.51 (0.32, 0.82)** |
|  | 60-74 years | **0.71 (0.52, 0.98)** | 0.76 (0.55, 1.06) | 1.11 (0.75, 1.64) | 1.13 (0.75, 1.69) |
|  | ≥ 75 years | 0.81 (0.52, 1.24) | 0.94 (0.60, 1.46) | 1.00 (0.58, 1.74) | 0.88 (0.50, 1.55) |
|  | Missing | N.E. | N.E. | N.E. | N.E. |
| Ethnicity/Race/Sex | NH Black Female | 1 | 1 | 1 | 1 |
|  | NH White Female | **0.46 (0.31, 0.67)** | **0.46 (0.32, 0.67)** | **2.40 (1.64, 3.53)** | **2.42 (1.64, 3.56)** |
|  | Hispanic Female | **3.11 (1.98, 4.86)** | **3.08 (1.96, 4.84)** | 1.38 (0.64, 2.99) | 1.36 (0.62, 2.97) |
|  | Other Female | 0.91 (0.53, 1.57) | 0.92 (0.53, 1.60) | **2.42 (1.32, 4.45)** | **2.47 (1.34, 4.57)** |
|  | NH White Male | 0.75 (0.53, 1.06) | 0.77 (0.55, 1.09) | **1.61 (1.04, 2.49)** | **1.59 (1.02, 2.47)** |
|  | NH Black Male | 0.77 (0.59, 1.02) | 0.79 (0.60, 1.04) | 0.70 (0.45, 1.09) | 0.72 (0.46, 1.13) |
|  | Hispanic Male | **2.92 (1.86, 4.58)** | **3.01 (1.90, 4.77)** | 1.94 (0.98, 3.85) | **2.04 (1.02, 4.08)** |
|  | Other Male | 0.99 (0.54, 1.80) | 0.99 (0.54, 1.81) | 1.82 (0.87, 3.84) | 1.87 (0.89, 3.94) |

Abbreviations: Adj. Adjusted; OR Odds Ratio; CI confidence interval; n, number; NH non-Hispanic.

We then disaggregated the data by sex, race and ethnicity. In terms of natural infection (**Supplemental Table 2a**), individuals between the ages of 45 to 74 were 22% less likely to have evidence of natural infection than the 18- to 29-year-old JHH ED patients. White individuals were 30% less likely to have been infected compared to Black individuals. Hispanic individuals had more than three times the burden of infection compared to non-Hispanic individuals, aOR 3.31 (95% CI 2.16, 5.07). After adjusting for age, and race/ethnicity, women had an increased odds for vaccination compared to men, aOR 1.35 (95% CI: 1.02, 1.80, **Supplemental Table 2b**). In comparison to patients aged 18-29, patients aged 45-59 years less likely to be vaccinated, aOR 0.50 (95% CI: 0.31, 0.80). Furthermore, White patients had more than twice the odds of vaccination compared to Black patients, aOR 2.26, 95% CI: 1.67, 3.07) There did not appear to be a significant difference in vaccination with respect to ethnicity, aOR1.25, 95% CI: 0.69, 2.29).

## Discussion

This study describes a method for distinguishing between SARS-CoV-2 vaccinated, naturally infected, and uninfected individuals using commercially available serologic assays when no previous vaccination or infection history is available. The algorithm utilized in this study indicates a 10-fold increase in seropositivity to SARS-CoV-2 infection in the Baltimore metropolitan area from April 2020 to March 2021. Furthermore, this study highlights disparities based on sex and race/ethnicity in SARS-CoV-2 prevalence and vaccine distribution within metropolitan Baltimore during the spring of 2021.

Our study follows the work of Suhandynata et al.^29^, in the ability to differentiate vaccinated from infected individuals based on antibody responses to the S1 and N proteins of SARS-CoV-2. In contrast to the Suhandynata et al. study which utilized chemiluminescent assays (Roche Elecsys Anti-SARS-CoV-2 S- and N-antibody)^29^, we applied more commonly available ELISA and LFA methods in our study. We further expanded their work by incorporating larger validation cohorts and applying the algorithm to population level surveillance. Our study also confirms previous reports of high burden of COVID-19 among the Baltimore Hispanic population^30^. Additionally, the discrepancies in vaccine uptake among racial and ethnic minority groups are clearly demonstrated. While recent data suggest that racial and ethnic gaps in vaccination have narrowed^10^, our data from early 2021 suggests that disparities in vaccination were present in the initial stages of the vaccine rollout. Surprisingly, despite prioritizing older Americans during the vaccine rollout^31^, patients older than 60 were as likely to be vaccinated as those between the ages of 18-44.

Our method demonstrated 100% sensitivity in identifying individuals who were fully vaccinated with both the Pfizer and Moderna vaccines. Further studies are needed to determine if other authorized vaccines have similar performance characteristics. One critical potential limitation to the use of anti-spike and anti-nucleocapsid antibody testing to differentiate naturally infected from vaccinated individuals is differential loss of antibody reactivity to these two targets. In a cohort of 3276 UK healthcare workers, Lumley et al. estimated that anti-nucleocapsid IgG antibodies exhibit a half-life of 85 days from the maximum titer (95% CI, 81-90)^32^. In contrast, the half-life of anti-spike IgG antibodies could not be measured, as 94% of healthcare workers did not exhibit significant loss during follow-up^32^. Additionally, anti-nucleocapsid antibody decline was more rapid in younger patients and those with milder symptoms. Thus, in the proposed algorithm, a proportion of naturally infected individuals will be misclassified as vaccinated as anti-nucleocapsid antibodies wane with time. This effect will be differential by age and initial symptomology. The time to seroreversion of spike and nucleocapsid antibodies in SARS-CoV-2 infected patients is significantly affected by both disease severity and assay platform. The effect of these variations should be considered when interpreting the results of serosurveillance studies.

Our study has several additional limitations. We did not test any individuals with known breakthrough infection (vaccinated then infected), nor could we distinguish between naturally infected individuals who were and were not subsequently vaccinated. Furthermore, a lack of seroreactivity occurred in a minority of naturally infected individuals. The lack of seroconversion in infected individuals has been observed in other studies, and occurs most frequently in individuals with asymptomatic infection ^33–35^. Using our testing algorithm, 16% of individuals with a previous positive RT-PCR test were seronegative by our algorithm. Similarly, Self et al. found that in a convenience sample of 156 mildly infected frontline healthcare personnel, 93.6% experienced a decline in antibody response and 28.2% seroreverted within 60 days^36^. These studies illustrate the difficulty of identifying infected persons several months after infection, especially in cases of mild infection. It should be noted that antibody reactivity is also dependent on the assay used, especially at 6 months after SARS-CoV-2 infection^37^.

Although the correlates of antibody protection for naturally infected individuals have not been well-established^38^, we demonstrated that a serosurvey can be performed to differentiate vaccinated, naturally infected and at-risk unexposed individuals in a population when vaccination or infection history is not available. This information provides evidence for targeted public health intervention in preparation for the continued spread of endemic SARS-CoV-2 infections.

## Data Availability

All data produced in the present study are available upon reasonable request to the authors

## Acknowledgements

The authors would like to acknowledge the staff and patients at the Johns Hopkins Hospital for making this research possible. We thank Morgan Keruly, Ethan Klock and Olivia Ajayi for technical assistance. We would also like to thank the generosity of the collective community of donors to the Johns Hopkins University School of Medicine and the Johns Hopkins Health System for COVID research.

## Figure Legends

**Supplemental Table 1:** Characteristics of Utilized Commercial SARS-CoV-2 Assays

| Manufacturer | Assay Name | Target antigen (recombinant) | Platform | Manufacturer's Interpretation |
| --- | --- | --- | --- | --- |
| Euroimmun, Lubeck, Germany | Anti-SARS-CoV-2 ELISA (IgG) | Spike-1 protein | Manual ELISA | Negative: S/C ratio <0.8<br>Borderline: S/C ratio ≥0.8 & <1.1<br>Positive: S/C ratio ≥1.1 |
| Hangzhou Biotest Biotech Co. Ltd., Hangzhou, China | CoronaCHEK™ COVID-19 IgG/IgM Rapid Test Cassette | Spike RBD | Lateral Flow Assay | IgG and IgM Positive: Three lines appear<br>IgG Positive: Control and IgG lines appear<br>IgM Positive: Control and IgM lines appear<br>Negative: One line in control region<br>Invalid: No control line |
| Bio-Rad Laboratories, Inc., Hercules, CA, USA | Platelia SARS-CoV-2 Total Ab assay | Nucleocapsid Protein | Manual ELISA | Negative: S/C ratio < 0.8<br>Equivocal: S/C ratio ≥ 0.8 & < 1.0<br>Positive: S/C ratio ≥ 1.0 |

Abbreviations: ELISA, enzyme-linked immunosorbent assay; S/C, signal to control; RBD, receptor binding domain.

**Supplemental Table 2a:**
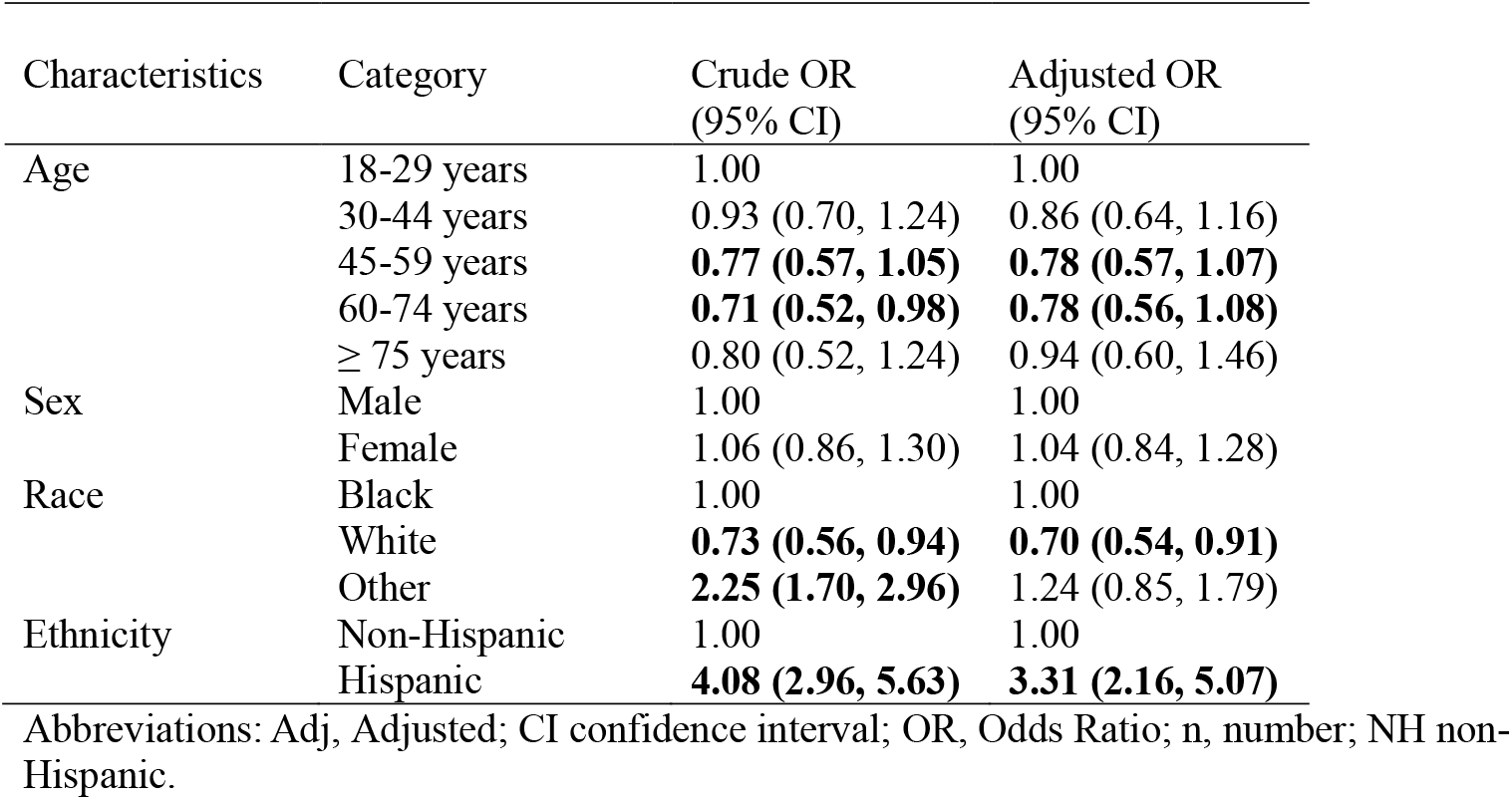
Demographic Characteristics Associated with Natural Infection among 2595 Emergency Department Patients, January –March 2021

| Characteristics | Category | Crude OR<br>(95% CI) | Adjusted OR<br>(95% CI) |
| --- | --- | --- | --- |
| Age | 18-29 years | 1.00 | 1.00 |
|  | 30-44 years | 0.93 (0.70, 1.24) | 0.86 (0.64, 1.16) |
|  | 45-59 years | **0.77 (0.57, 1.05)** | **0.78 (0.57, 1.07)** |
|  | 60-74 years | **0.71 (0.52, 0.98)** | **0.78 (0.56, 1.08)** |
|  | ≥ 75 years | 0.80 (0.52, 1.24) | 0.94 (0.60, 1.46) |
| Sex | Male | 1.00 | 1.00 |
|  | Female | 1.06 (0.86, 1.30) | 1.04 (0.84, 1.28) |
| Race | Black | 1.00 | 1.00 |
|  | White | **0.73 (0.56, 0.94)** | **0.70 (0.54, 0.91)** |
|  | Other | **2.25 (1.70, 2.96)** | 1.24 (0.85, 1.79) |
| Ethnicity | Non-Hispanic | 1.00 | 1.00 |
|  | Hispanic | **4.08 (2.96, 5.63)** | **3.31 (2.16, 5.07)** |

Abbreviations: Adj, Adjusted; CI confidence interval; OR, Odds Ratio; n, number; NH non-Hispanic.

**Supplemental Table 2b:** Demographic Characteristics Associated with Evidence of Vaccination among 2380 Emergency Department Patients, January –March 2021

| Characteristics | Category | Crude OR<br>(95% CI) | Adjusted OR<br>(95% CI) |
| --- | --- | --- | --- |
| Age | 18-29 years | 1.00 | 1.00 |
|  | 30-44 years | 1.03 (0.70, 1.51) | 0.98 (0.67, 1.45) |
|  | 45-59 years | **0.48 (0.30, 0.77)** | **0.50 (0.31, 0.80)** |
|  | 60-74 years | 1.05 (0.70, 1.56) | 1.09 (0.73, 1.65) |
|  | ≥ 75 years | 0.96 (0.55, 1.68) | 0.86 (0.49, 1.52) |
| Sex | Male | 1.00 | 1.00 |
|  | Female | **1.36 (1.03, 1.80)** | **1.35 (1.02, 1.80)** |
| Race | Black | 1.00 | 1.00 |
|  | White | **2.30 (1.71, 3.11)** | **2.26 (1.67, 3.07)** |
|  | Other | **2.61 (1.73, 3.91)** | **2.42 (1.51, 3.88)** |
| Ethnicity | Non-Hispanic | 1.00 | 1.00 |
|  | Hispanic | **1.88 (1.13, 3.13)** | 1.25 (0.69, 2.29) |

Abbreviations: Adj, Adjusted; CI confidence interval; OR, Odds Ratio; n, number; NH non-Hispanic.

**Supplemental Figure 1a.**
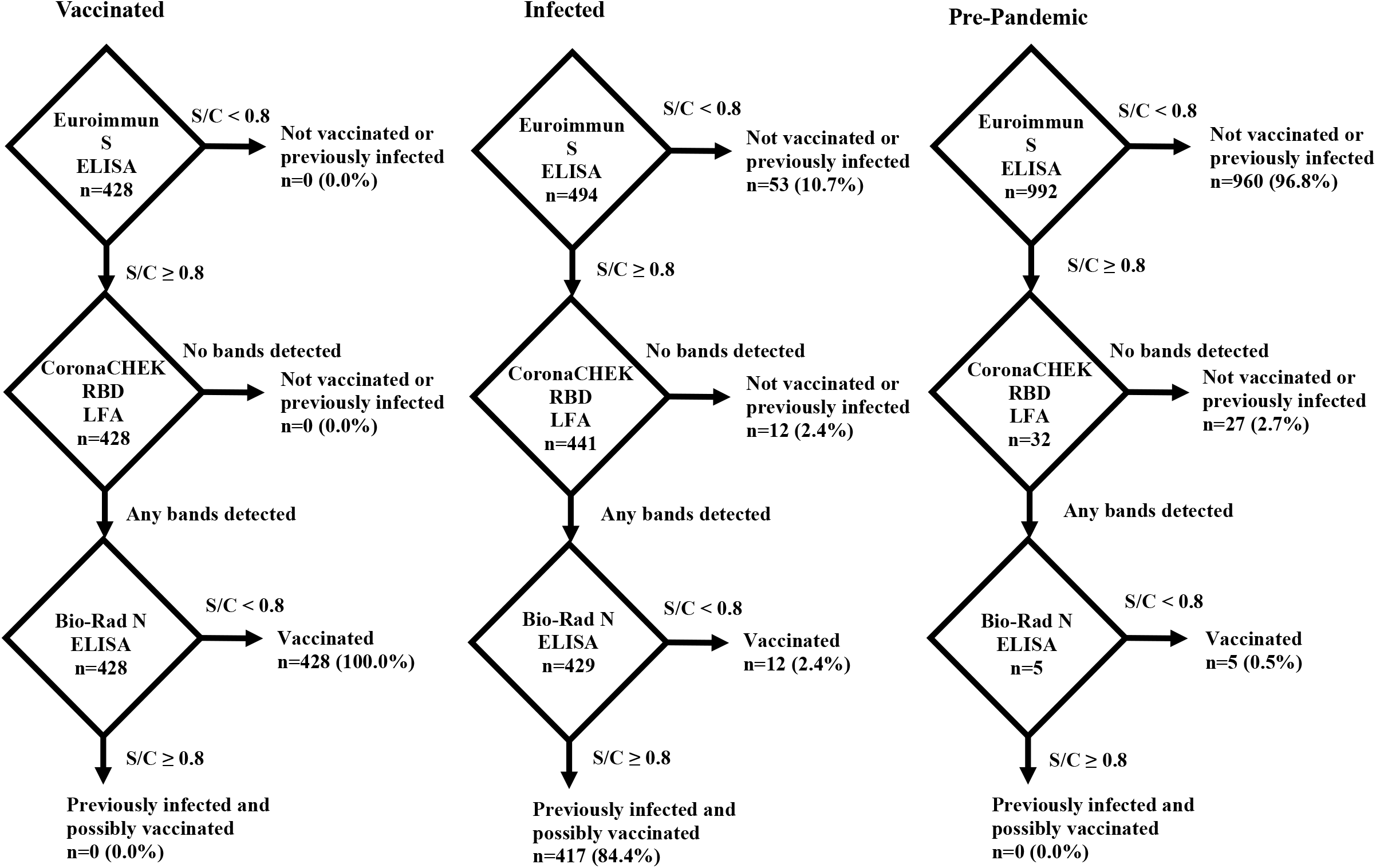
Testing algorithm results on samples from known vaccinated, naturally infected and pre-pandemic samples.

**Supplemental Figure 1b.**
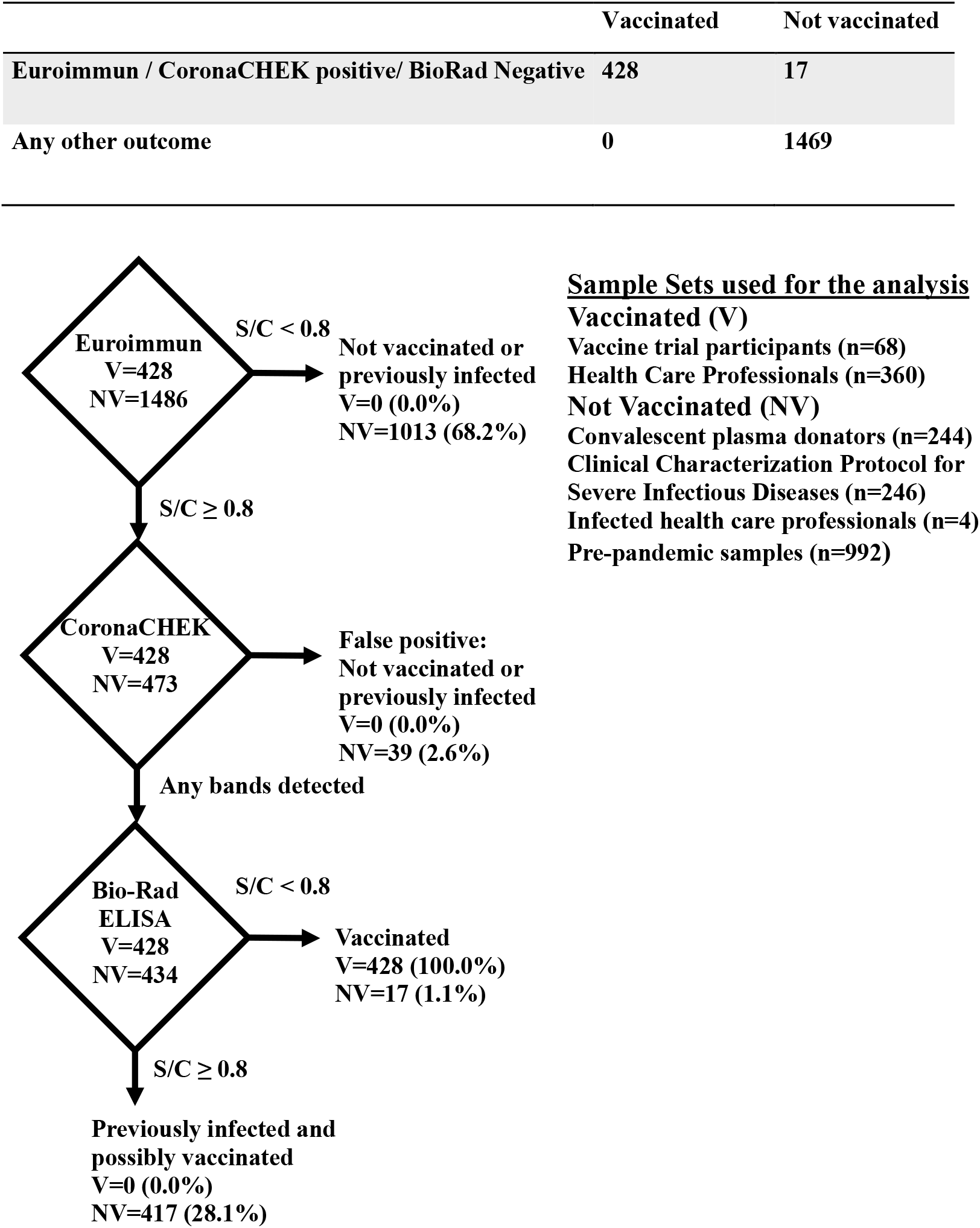
Determination of sensitivity and specificity of vaccinated state in testing algorithm.

**Supplemental Figure 1c.**
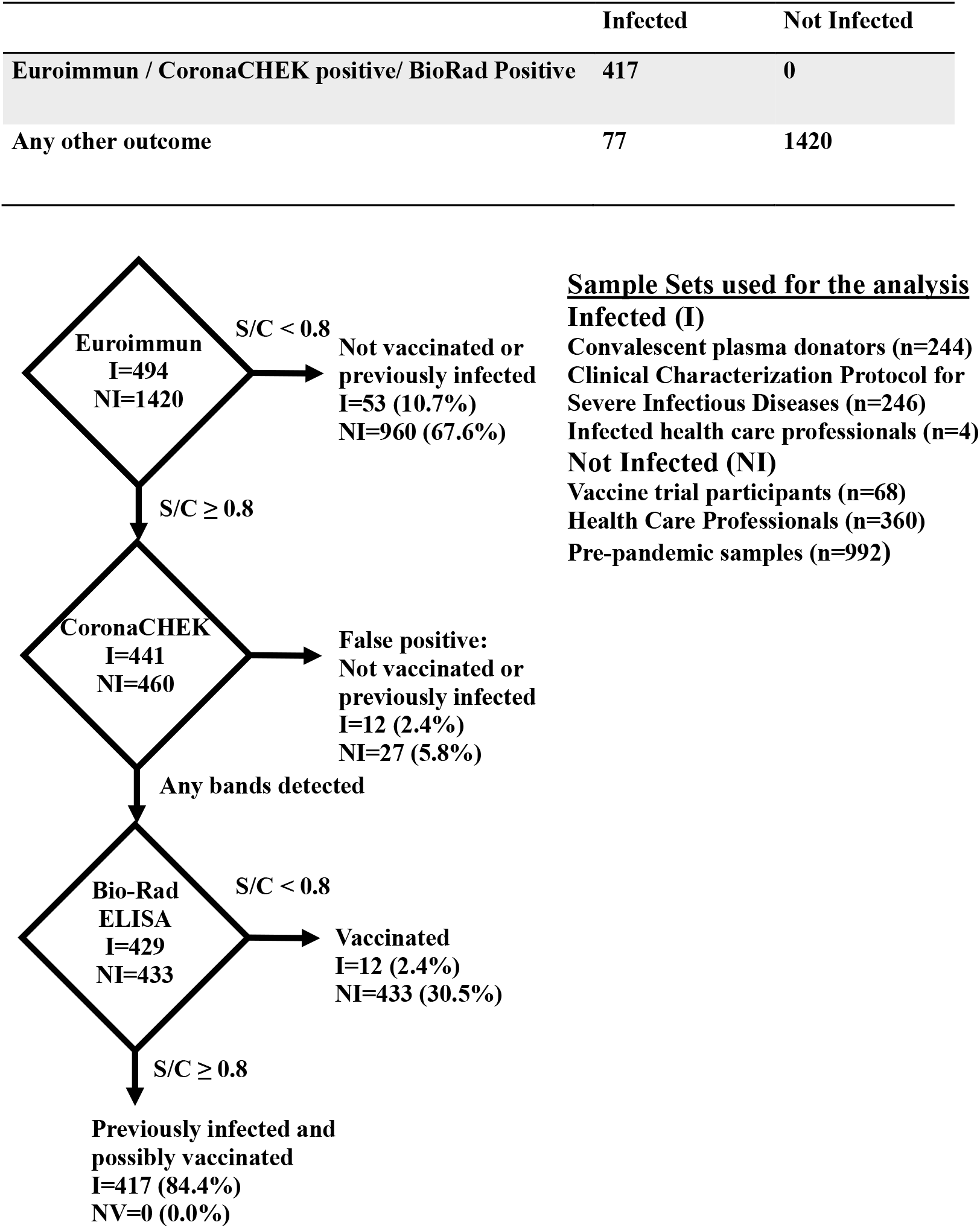
Determination of sensitivity and specificity of naturally infected state in testing algorithm.

**Supplemental Figure 2a.**
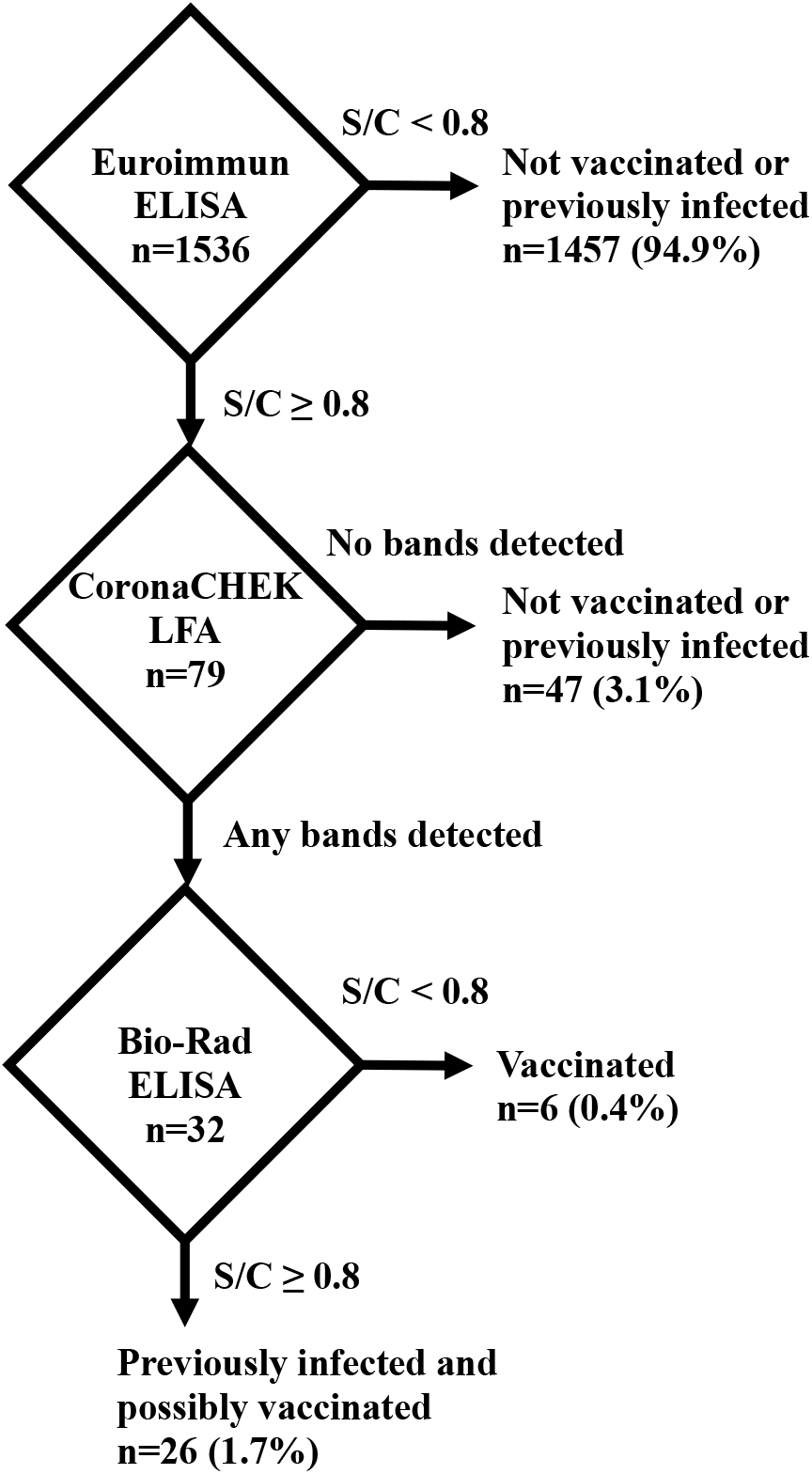
Testing algorithm results for ED samples collected in 2020.

**Supplemental Figure 2b.**
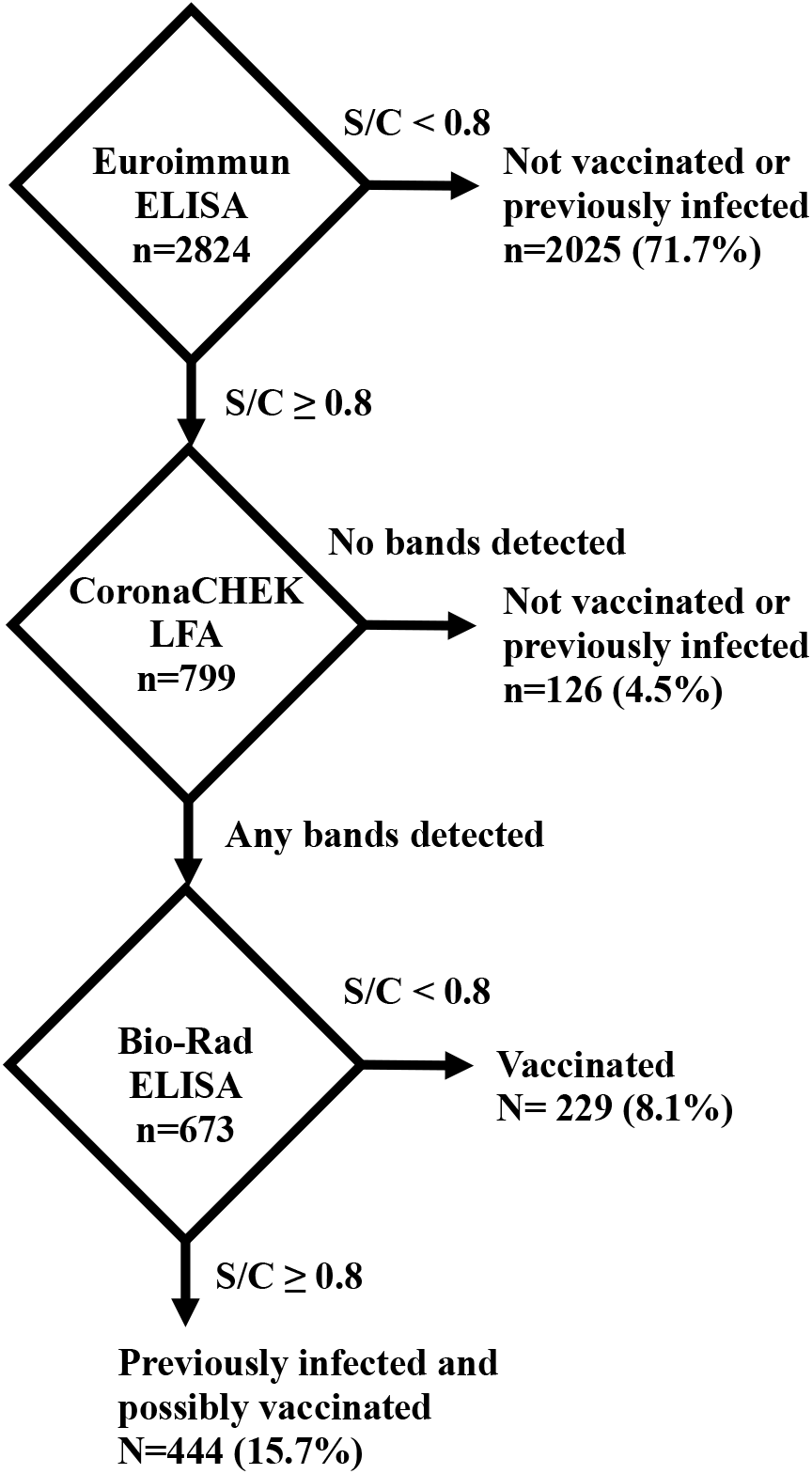
Testing algorithm results for ED samples collected in 2021.

**Supplemental Figure 3a.**
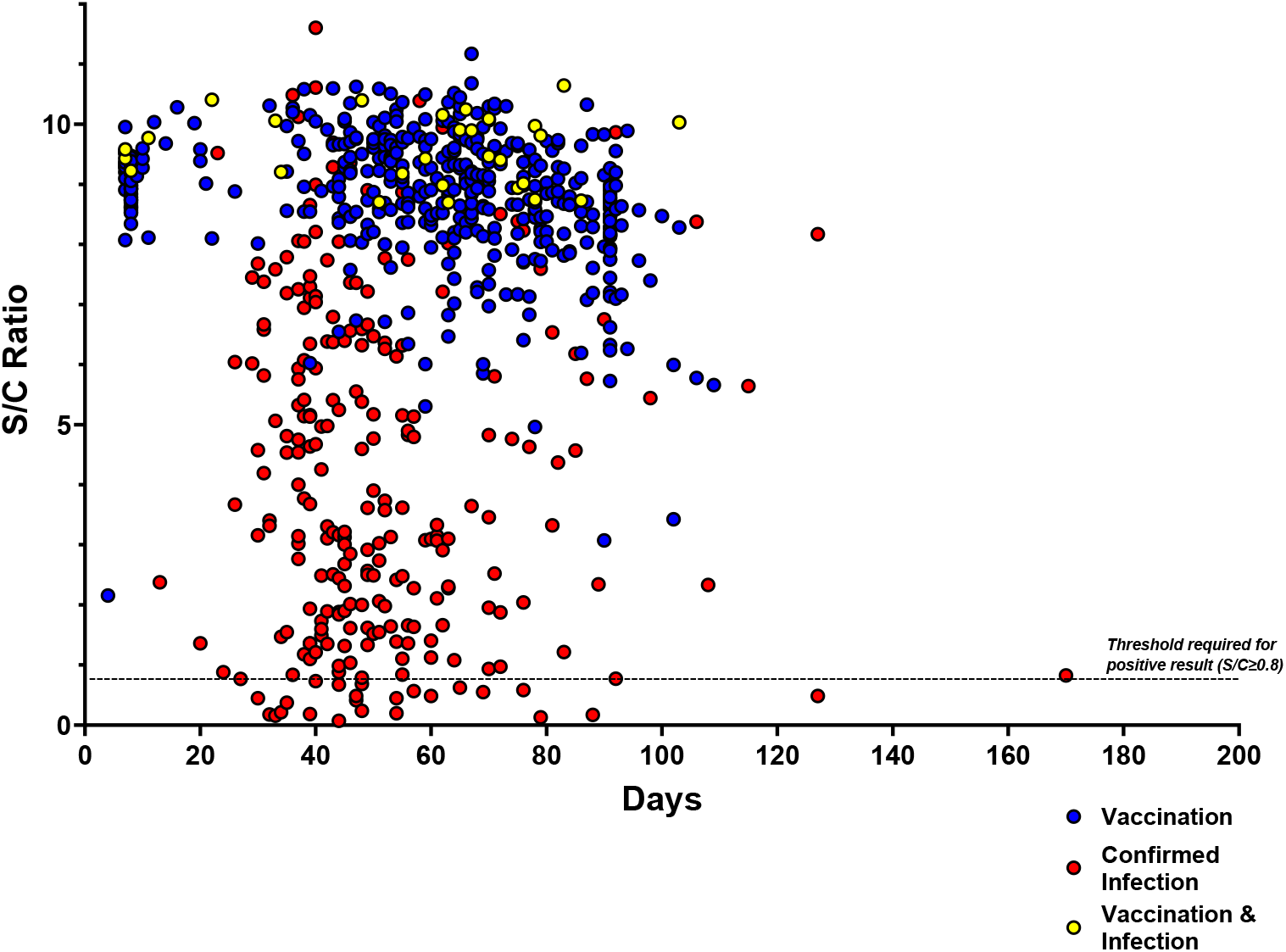
Euroimmun S/C values of antibody response against spike protein by days post-second dose or confirmed infection. Euroimmun index values were plotted against time since receipt of the second dose of a vaccine (vaccinated samples) or time since confirmed SARS-CoV-2 PCR-positive infection (infected samples and vaccinated/infected samples).

**Supplemental Figure 3b.**
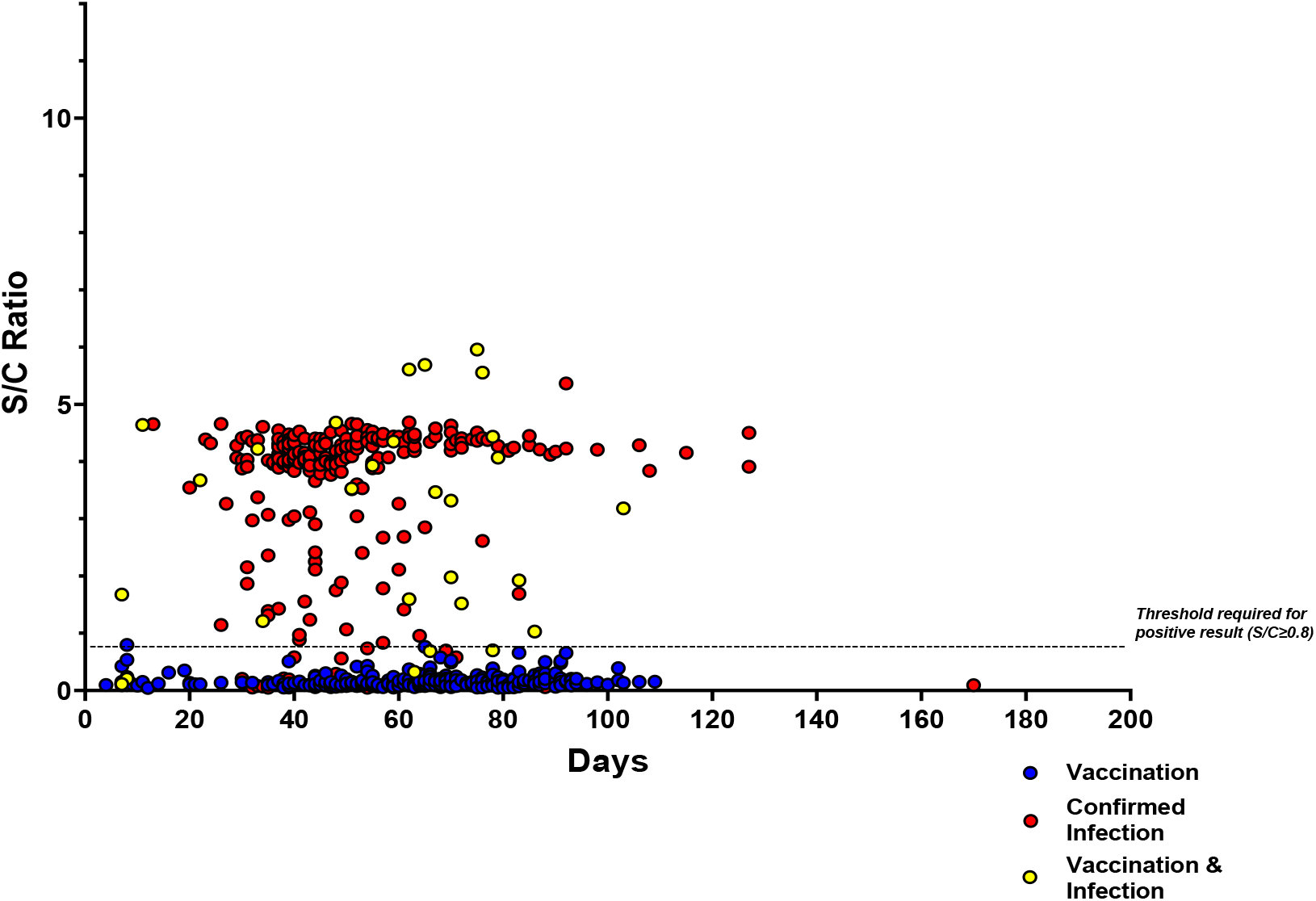
Bio-Rad S/C values of antibody response against nucleocapsid by days post-second dose or confirmed infection. Bio-Rad index values were plotted against time since receipt of the second dose of a vaccine (vaccinated samples) or time since confirmed SARS-CoV-2 PCR-positive infection (infected samples and vaccinated/infected samples).

